# Clinical Study on Human Gut Microbiota and Short-Chain Fatty Acids in Response to Dietary Interventions with high resistance Potato starch

**DOI:** 10.64898/2026.07.15.26358131

**Authors:** Gunvant Yeola, Almas yaser khan, Amruta Bhagwan More, Amol Hartalkar, Rakesh Shantanu Hingane, Ruby Dubey, Ritika Singhvi, Kiran Khatau

**Affiliations:** sponsor :-Lodaat Pharma 1415 1415 WEST 22ND STREET, TOWER FLOOR, Oak Brook, IL 60523, US; CRO :- Target Institute of Medical Education & Research Pvt Ltd, Mumbai; Prabhankur Wellness Clinic, Ground Floor, Department of OPD, OPD No. 1, DSK SUNDARBAN PHASE-1, Shop No. 5, near Amanora Park Town, Phase 01, Main Gate, Hadapsar, Pune, Maharashtra; Yashasvi Hospital Hingane Nivas, Kale Borate Nagar Road, Pune, MH-411028; Dr. D. Y. Patil College of Ayurved and Research Centre Pune

## Abstract

**Background and Objective:** To investigate the efficacy of a new, proprietary high-resistance potato starch as a prebiotic in comparison to inulin and a control.

**Methods:** In this prospective study, an intervention of 9 g of resistant potato starch (RPS, Potatodaat^®^), inulin, or accessible corn starch was given to participants with mild to moderate indigestion for 30 days. Short chain fatty acid (SCFA) levels, changes to the gut microbiome, and changes in clinical symptoms of indigestion were assessed as primary outcomes.

**Results:** Subjects in the RPS (n = 22), inulin (n = 23), and accessible corn starch (n = 22) groups demonstrated similarity in age, sex, and baseline parameters. At 30 days, the groups experienced 21.1%, 6.29%, and 9.15% increases in stool butyrate, respectively. Clinical symptoms like indigestion and flatulence showed greater improvement with the use of RPS compared to the other two groups.

**Conclusion:** Consumption of high resistance potato starch helps improve stool SCFA levels along with clinical symptoms related to digestion, showing its better prebiotic potential as compared to inulin and accessible corn starch. The new high-resistance potato starch can be considered an effective replacement for inulin.

## Introduction

The gut microbiome is considered an integral part of human health. Research shows a strong relationship between gut microbiome and immune, skin and brain health in addition to digestive health. Kenneth J. O’Riordan, Gerard M. Moloney, Lily Keane, Gerard Clarke, John F. Cryan,

The gut microbiota-immune-brain axis: Therapeutic implications, Cell Reports Medicine, Volume 6, Issue 3,2025, 101982. The gut microbiome spectrum consists of various microorganisms like bacteria, archaea, viruses and fungi (mostly yeasts). Over 35 trillion bacteria live in the human gastrointestinal (GI) tract, the vast majority of which reside in the colon^1^. Gut microbiota is highly active and deeply involved in the mucosal immune system and the maintenance of intestinal homeostasis. Intestinal microbiota is divided into three categories considering their role in the host^2-5^. The first category is physiological bacteria which shows a favorable symbiotic relationship with the host. These mostly anaerobic bacteria attach to deep mucosal epithelial cells. They are the dominant microbiota of the intestine and play key roles in nutrition and immune regulation (*e.g*., *Bifidobacterium, Bacteroides* and *Peptococcus).* The second category is conditional pathogens (*e.g*., *Enterococcus* and *Enterobacter)* that inhabit the host. They are harmless when intestinal microecological balance is maintained but can be harmful to humans under certain conditions. The third category contains mostly pathogens (*e.g*., *Proteus* and *Pseudomonas^2-5^*). These bacteria typically cause disease in the host.

A variety of factors influence the composition and function of microbiota in the human gut including genetics, diet, age, use of antibiotics, stress, environmental factors and psychological factors. Disturbed microbiome composition and dysfunction are linked to various conditions and diseases like cancer, obesity, metabolic disease, diabetes, allergies, and depression. Barkha Madhogaria, Priyanka Bhowmik, Atreyee Kundu, Correlation between human gut microbiome and diseases, Infectious Medicine, Volume 1, Issue 3, 2022, Pages 180-191, ISSN 2772-431X, https://doi.org/10.1016/j.imj.2022.08.004.

A growing body of research is focusing on targeted interventions to modulate and improve the gut microbiome through dietary supplementation. Prebiotics are plant fibers that support the growth of healthy bacteria in the gut by acting as a food source. Resistant starches are such an example of prebiotics. Enzymes early in the digestive tract are largely unable to digest resistant starches, allowing them to pass to the colon where they undergo anaerobic fermentation by bacteria residing there. This degradation by bacteria releases intermediates, such as acetate and lactate, that cross-feed with bacteria that produce short chain fatty acids, such as butyrate. Butyrate is the most important Short Chain Fatty Acid which helps in promoting satiety, blood sugar management, and provides protection from endothelial dysfunction. Scientific studies have also found that these Prebiotics also help in the management of blood sugar. These prebiotics can help the food to ferment and digest faster. These also help to keep the inner lining of the intestine healthy and to absorb certain minerals like calcium and magnesium.

Inulin, a commonly used prebiotic has been reported to cause Nausea, bloating, and flatulence and can cause side effects in patients with inflammatory bowel disease (IBD) or allergies. Sheng W, Ji G, Zhang L. Immunomodulatory effects of inulin and its intestinal metabolites. Front Immunol. 2023 Aug 10;14:1224092. doi: 10.3389/fimmu.2023.1224092. PMID: 37638034; PMCID: PMC10449545.

Since low quantities of fructo-oligosaccharides and galacto-oligosaccharides naturally exist in foods, scientists are attempting to produce prebiotics on an industrial scale. Considering the health benefits of prebiotics and their safety, as well as their production and storage advantages compared to probiotics, they seem to be fascinating candidates for promoting human health condition as a replacement or in association with probiotics. (Davani-Davari D, Negahdaripour M, Karimzadeh I, Seifan M, Mohkam M, Masoumi SJ, Berenjian A, Ghasemi Y. Prebiotics: Definition, Types, Sources, Mechanisms, and Clinical Applications. Foods. 2019 Mar 9;8(3):92. doi: 10.3390/foods8030092. PMID: 30857316; PMCID: PMC6463098. to be added)

Resistant starch has the potential to serve as an equally efficacious alternative to inulin with fewer side effects. Resistant starch can be classified in five categories. (Birt DF, Boylston T, Hendrich S, Jane JL, Hollis J, Li L, McClelland J, Moore S, Phillips GJ, Rowling M, Schalinske K, Scott MP, Whitley EM. Resistant starch: promise for improving human health. Adv Nutr. 2013 Nov 6;4(6):587-601. doi: 10.3945/an.113.004325. PMID: 24228189; PMCID: PMC3823506) RSI refers to physically inaccessible starches as found in coarsely ground or whole-kernel grains. RSII refers to granular starches such as high-amylose maize starches and raw banana starch. RSIII is retrograded starch from heated and cooled starchy foods, and RSIV refers to chemically modified starches such as distarch phosphate and octenyl succinate starch. RSV are amylose-lipid complexes that decrease starch accessibility. In 2018, the USFDA announced that RS2 would be included as a source of dietary fiber. (Review of the Scientific Evidence on the Physiological Effects of Certain Non-Digestible Carbohydrates, Office of Nutrition and Food Labeling Center for Food Safety and Applied Nutrition Food and Drug Administration U.S. Department of Health and Human Services, June 2018)

This study evaluates a novel prebiotic resistant starch (RS2) sourced from potatoes as a dietary intervention. The material contains up to 65% resistant starch. Looking at the activities of Potatodaat^®^, a clinical study titled “Evaluation of efficacy and safety of a proprietary high resistance Potato starch-Potatodaat^®^ as a prebiotic by SCFA analysis and Gut Microbiota in Healthy individuals – A Double Blind, Randomized, Active Controlled, Multicenter, Comparative, Interventional, Prospective, Clinical Study” was conducted.

## Materials and Methods

### Design

The study was a double-blinded, randomized, active-controlled, multicenter, comparative, interventional, prospective, three-arm clinical study. The study was conducted at Dr. D. Y. Patil College Of Ayurved And Research Centre Pimpri, Pune, Maharashtra, India and Prabhakar Wellness Clinic, Hadapsar, Pune, Maharashtra, India. The study was approved by the institutional and independent ethics committee of the two study sites and was registered with CTRI wide registration no. CTRI/2022/12/048064.

### Participants (Inclusion & Exclusion Criteria)

Healthy male and female participants of age between 20 and 65 years (both inclusive) willing to provide written informed consent and follow the study protocol were considered for screening. Healthy individuals were considered as those who did not have any medical or surgical conditions for which they had to take immediate or continuous medical or surgical treatment. Participants with BMI between 21 kg/m^2^ to <28 kg/m^2^ were included. Participants having mild to moderate indigestion (occasional mild to moderate bothersome postprandial fullness and or early satiation, epigastric pain) were recruited in the study.

Exclusion criteria in the study included pregnant and lactating females, confirmed diagnosis of COVID-19, and recent diagnosis (<1 month) of COVID-19. Additionally, individuals with known cases of diabetes, those of immunocompromised status (as in HIV, Hepatitis, Tuberculosis, and Cancer), or those undergoing steroid and/or immunosuppressive therapy were excluded from the study. Participants who were currently part of any other clinical study or had participated in any other clinical study in the 3 months prior to screening in the present study were excluded. Participants with a history of allergy to potato starch, inulin, and accessible corn starch were also excluded. Lastly, any other conditions which, in the opinion of the investigators (like ulcerative colitis, gastric ulcers, etc), made the participant unsuitable for enrollment or that could interfere in adherence to the study protocol were excluded from the study.

### Intervention, Sample Size and Randomization

Participants were randomized to one of three groups i.e. proprietary high resistance potato starch (RPS, Potatodaat^®^, Lodaat LLC) group, inulin supplement group, or accessible corn starch group in 1:1:1 ratio. Participants were asked to consume the given study product in a dose of 9 gm daily after breakfast with water for 30 days. Participants could mix the given study product with either cool/room temp water or with any other cool/room temp liquid and consume for 30 days.

A total of 75 participants were screened in the study of which 72 participants were recruited, 24 in each of the 3 groups. There subjects were lost to follow up (2 subjects in corn starch group, 2 in the RPS group, and 1 in the inulin group). A total of 67 participants completed the study, 22 in corn starch group, 23 in the Inulin group and 22 in the RPS group. None of the study participants dropped out due to adverse events related to the study product or procedure. All participants who completed the study were evaluated for efficacy assessment while all participants who took even a single dose of the study product were considered for safety assessment.

Randomization of subjects was done through a computer-generated randomization list which ensured distribution of participants in ratio of 1:1:1 in the three study groups. Packing and dispensing of study products was done to ensure that neither the participant nor the investigator knew about the group assigned to the participants. Sealed envelopes with the study groups were kept and investigators were asked to open the same only in case of any adverse events which may be related to the study product.

### Study duration and visits

The total duration of the study was 30 days (the duration for which the study products were consumed by the participants). After Screening Visit (Up to Day -3) participants were asked to come for a randomization visit which was considered as the Baseline Visit (Day 0) and thereafter 15 days and 30 days. A window period of 3 days was allowed in case of delayed follow up.

### Primary outcomes

The primary outcomes included comparative assessment of changes in stool short-chain fatty acids (SCFA) including butyrate, acetate, propionate, and total SCFA levels between the three groups over a period of 30 days (Time Points: Day 0, Day 15 and Day 30). Additional primary outcomes were to a comparative assessment of change in diversity and abundance of gut micro-biota between among the groups over a period of 30 days (Time Points: Day 0 and Day 30). Gut microbiota studied included *Ruminococcus bromii, Clostridium chartatabidum,* and *Eubacterium rectale*.

### Secondary outcomes

Secondary outcomes included comparative assessment of changes in digestive symptoms like indigestion, appetite, bowel habits, constipation, flatulence, and fullness of abdomen on a graded scale over a period of 30 days between the three groups. Other secondary outcome included comparative change in consistency of stool on Bristol Stool Scale between the three groups over a period of 30 days. Along with this efficacy assessment, a safety assessment was also done by evaluating tolerability of study products through assessment of Adverse Events. Global assessment of overall change as per the participant as well as by the investigator was done. Comparative assessment of pre-intervention & post-intervention stool routine and microscopic between the three groups was also done.

### Study Methodology

After ethics committee’s approval and subsequent registration of the study on CTRI, screening of participants for possible recruitment was completed. Healthy male and female participants of age between 20 and 65 years (both inclusive) were screened as per the inclusion & exclusion criteria. On screening visit (Day -3), a written informed consent was obtained from participants for their participation in the study. Participant’s demographic details and medical history were noted in the CRF. Participants underwent general and systemic examinations including vitals. Participant’s stool routine and microscopic were done. Participants were asked to refrain from any Nutraceutical, Ayurvedic, homeopathic, herbal etc. medicines. Participants were called on baseline visit (day 0). On baseline visit (Day 0), participants were recruited if he/she met all the inclusion criteria. As per computer generated randomization list, participants were randomized to one of the three groups. Participants were asked to take the assigned study product in a dose of 9 gm daily after food for a period of 30 days.

On baseline visit, participants were provided with stool sample collection kit and detailed instructions/training on stool collection were given. Next morning, participants were asked to collect stool in the given tube for analysis of stool short-chain fatty acids (SCFA) and gut microbiome and submit sample to study site. Participant’s stool sample for analysis of stool short-chain fatty acids (SCFA) were repeated on day 15 and on day 30. Participant’s stool sample for analysis of gut microbiome was repeated on day 30.

### Clinical Symptom Assessment

On baseline visit, day 15 and day 30, participants were evaluated for symptoms like indigestion (on a 0 to 100 scale, where 0= Very poor digestion and 100= excellent digestion), appetite (on a 0 to 100 scale, where 0= Very poor appetite and 100= excellent appetite.), Bowel habits (on a 0 to 100 scale, where 0= Very poor bowel movements and 100= excellent bowel movements), constipation (mild - Occasional or intermittent symptoms; occasional use of stool softeners, laxatives, dietary modification, or enema, moderate-Persistent symptoms with regular use of laxatives or enemas, Severe-manual evacuation indicated) flatulence (on a 0 to 100 scale, where 0= No flatulence and 100= maximum flatulence) and fullness of abdomen (on a 0 to 100 scale, where 0= No fullness of abdomen and 100= maximum fullness of abdomen). Participants would undergo general and systemic examinations including vitals. Participants’ consistency of stool was evaluated on Bristol stool scale. Study product compliance was evaluated by enquiring about any missed dosage.

On final follow up visit (Day 30), participant’s global evaluation for overall change and Investigator’s global evaluation for overall change were done. The tolerability of the study products was assessed by the investigator and participants. All participants were closely monitored for any adverse events/ adverse drug reactions from baseline visit till the end of the study. Participant’s stool routine and microscopic were done at the end of 30 days. Participants were asked to stop the study product after completion of 30 days and were advised to take investigator’s advice for further treatment.

### Adverse effects and safety assessment

Evaluation of adverse events/adverse drug reactions at every follow-up visit and establishment of their relationship with the study product was assessed. The tolerability of the study product was evaluated by following the safety grades as “1” for excellent overall safety (no adverse event(s) reported), “2” for good overall safety (mild adverse event(s) reported which subside with or without medication), “3” for fair overall safety (moderate to severe adverse event(s) reported which subside with or without medication and do not necessitate stoppage of study products), and “4” for poor overall safety (serious adverse event(s) which necessitate stoppage of study). Safety was assessed by clinical review of all safety parameters, including the reported adverse events if any, vital signs including allergic reactions, etc.

### Statistical Methods

The data obtained in the studies were subjected to tests of significance using appropriate statistics. The data on discrete variables are represented as actual frequencies, i.e., n (%). The data on continuous variables are represented as mean±SD. GraphPad InStat Version 3.6 software was used for statistical analysis of data. A p-value of <0.05 was considered significant. Participants who completed the study as per the protocol were considered as “per-protocol population” and participants who took at least one dose of the study drug were considered as “safety population” and were evaluated accordingly.

## Results

### Study Population

A total of 75 participants were screened in the study of which 72 participants were recruited, 24 in each of the 3 groups. Five subjects were lost to follow up (2 subjects each in corn starch group & RPS group and 1 in inulin group). A total of 67 participants completed the study, 22 in corn starch group, 23 in Inulin group and 22 in RPS group. None of the study participants dropped out due to adverse events related to the study product or procedure. All participants who completed the study were evaluated for efficacy assessment while all participants who took even a single dose of the study product were considered for safety assessment. **Refer Figure 1**.

**Figure 1:**
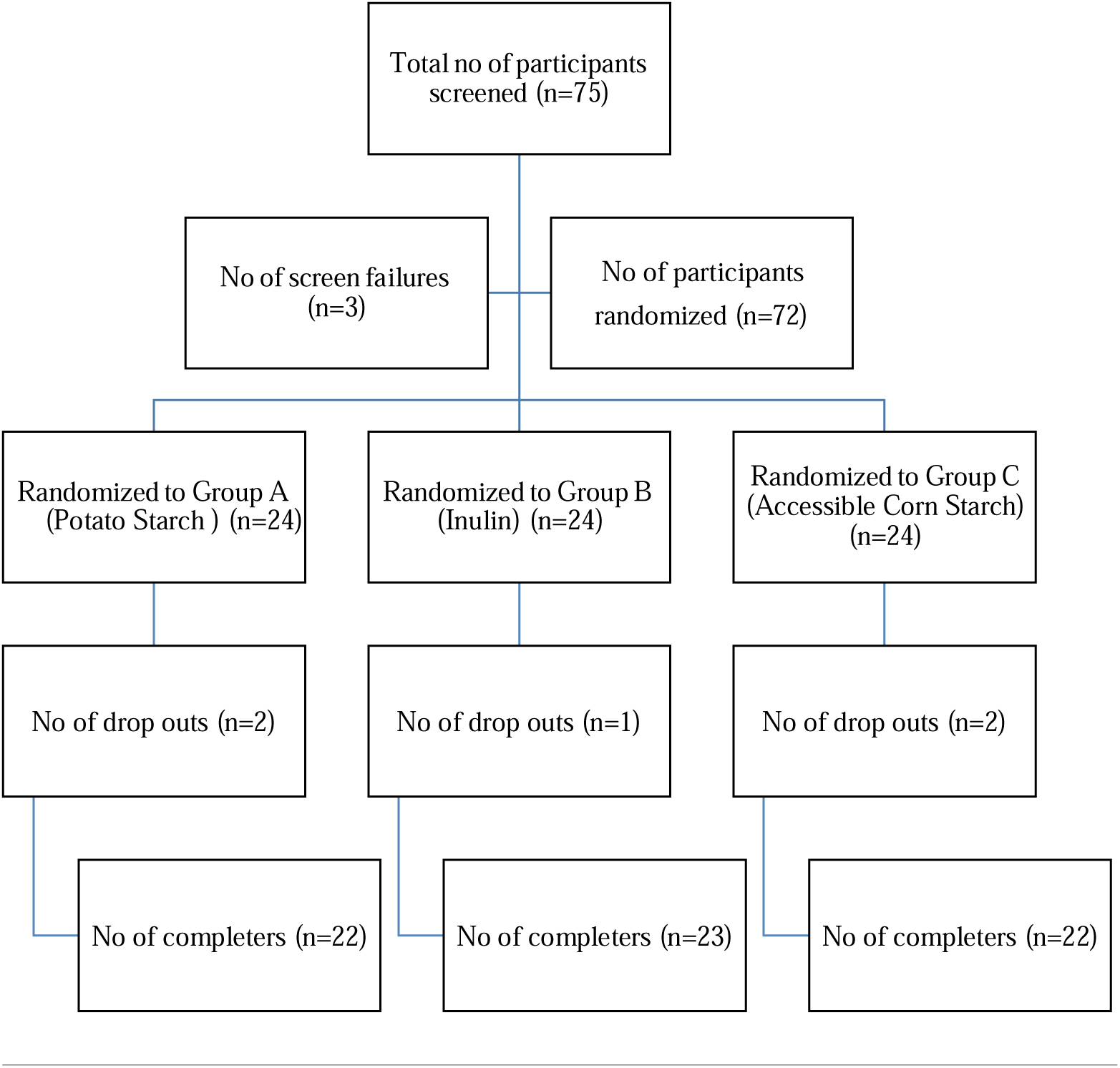
Participant Details (Study consort)

### Demographic details

There were a total of 12 males (54.54%) and 10 females (45.45%) in corn starch group while the male and females numbers were 13 (56.52%) and 10 (43.47%) in Inulin group respectively and that in RPS group was 10 (45.45%) and 12 (54.54%) respectively. The mean age of participants in corn starch group was 32.63 ±11.68, while that in inulin group it was 28.65 ±7.70 and that in RPS group it was 30.87 ±8.72 years. There was no significant difference between the mean age of participants between the three groups. The mean body weight of participants in corn starch group was 63.00 ±7.61 kg and the BMI was 23.92 ±1.71. The weight and BMI in inulin group were observed to be 61.63 ±5.77 kg and 23.62 ±1.54 kg respectively while in the RPS group it was 64.04 ±8.32 kg and 24.36 ±1.77 respectively. Analysis between the groups showed non-significant difference between the three groups. Stool pH at baseline in corn starch group was observed to be 6.63 ±0.23 while in the inulin group it was 6.61 ±0.33 and in the RPS group it was 6.57 ±0.45, showing no significant difference. **Refer table 1** for details on demography. Distribution of participants as per the diet that they consumed is detailed in **table 2**.

**Table 1:** Baseline demography in the 3 study groups.

| Groups | Age | Gender | Weight | BMI | Stool pH |
| --- | --- | --- | --- | --- | --- |
| Corn Starch Group (n=22) | 32.63<br>± 11.68 | Male-12<br>(54.54%)<br>Females-10<br>(45.45%) | 63.00<br>± 7.61 | 23.92<br>± 1.71 | 6.63<br>± 0.23 |
| Inulin Group (n=23) | 28.65<br>± 7.70 | Male -13<br>(56.52%)<br>Females -10<br>(43.47%) | 61.63<br>± 5.77 | 23.62<br>± 1.54 | 6.61<br>± 0.33 |
| Potato starch Group (n=22) | 30.87<br>± 8.72 | Male -10<br>(45.45%)<br>Females -12<br>(54.54%) | 64.04<br>± 8.32 | 24.36<br>± 1.77 | 6.57<br>± 0.45 |
| P value between the groups | p>0.05 (NS) | p>0.05 (NS) | p>0.05 (NS) | p>0.05 (NS) | p>0.05 (NS) |

**Table 2:** Assessment of diet pattern of participants.

| Diet Type | Corn Starch Group (n=22) | Inulin Group (n=23) | Potato Starch Group (n=22) |
| --- | --- | --- | --- |
| Vegetarian foods including dairy products | 5 (22.72%) | 8 (34.78%) | 6 (27.27%) |
| Vegetarian foods excluding dairy products (Vegan) | 1 (4.54%) | 0 | 1(4.54%) |
| Mixed diet (Meat/Chicken/Eggs/Diaries & vegetables) | 16 (72.72%) | 15 (65.21%) | 15 (68.18%) |
| Regular Salad – Fruits/vegetable (Fiber rich food) eaters | 10 (45.45%) | 11 (47.82%) | 10 (45.45%) |
| P value between the groups | p>0.05 (NS) |  |  |

## Primary outcomes

### Assessment of change in stool acetate, propionate and butyrate levels

#### Changes in stool acetate levels

The baseline stool acetate levels in the corn starch group were observed to be 13.01 ±7.14 which showed a significant increase to 16.43 ±10.30 (p=0.04) at the end of 15 days and further to 17.65 ±12.09 (p=0.02). The levels of stool acetate levels in inulin group at baseline was observed to be 14.94 ±6.73 which increased significantly to 18.44 ±9.83 (p=0.02) at the end of 15 days and further to 22.20 ±12.46 (p=0.01) at the end of 30 days. In the RPS group, the stool acetate levels at baseline visit was 14.25 ±8.72 which showed a significant increase to a level of 18.97 ±10.79 (p=0.01) at the end of 15 days and further to 22.88 ±14.12 (p=0.01) at the end of 30 days. Analysis between the three groups showed that the increase in stool acetate levels were significantly higher in RPS group as compared to inulin and corn starch groups at 30 days (p=0.01). The percentage change in stool acetate levels from baseline to 15 days in Corn starch group were observed to be 26.28% while at 30 days it was observed to be 35.66%. The changes in stool acetate levels in inulin group were observed to be 23.42% at 15 days and 48.59 % at the end of 30 days. The levels increased by 33.12% at the end of 15 days and 60.56% at the end of 30 days in RPS group. **Refer Table 3 & 4.**

**Table 3:**
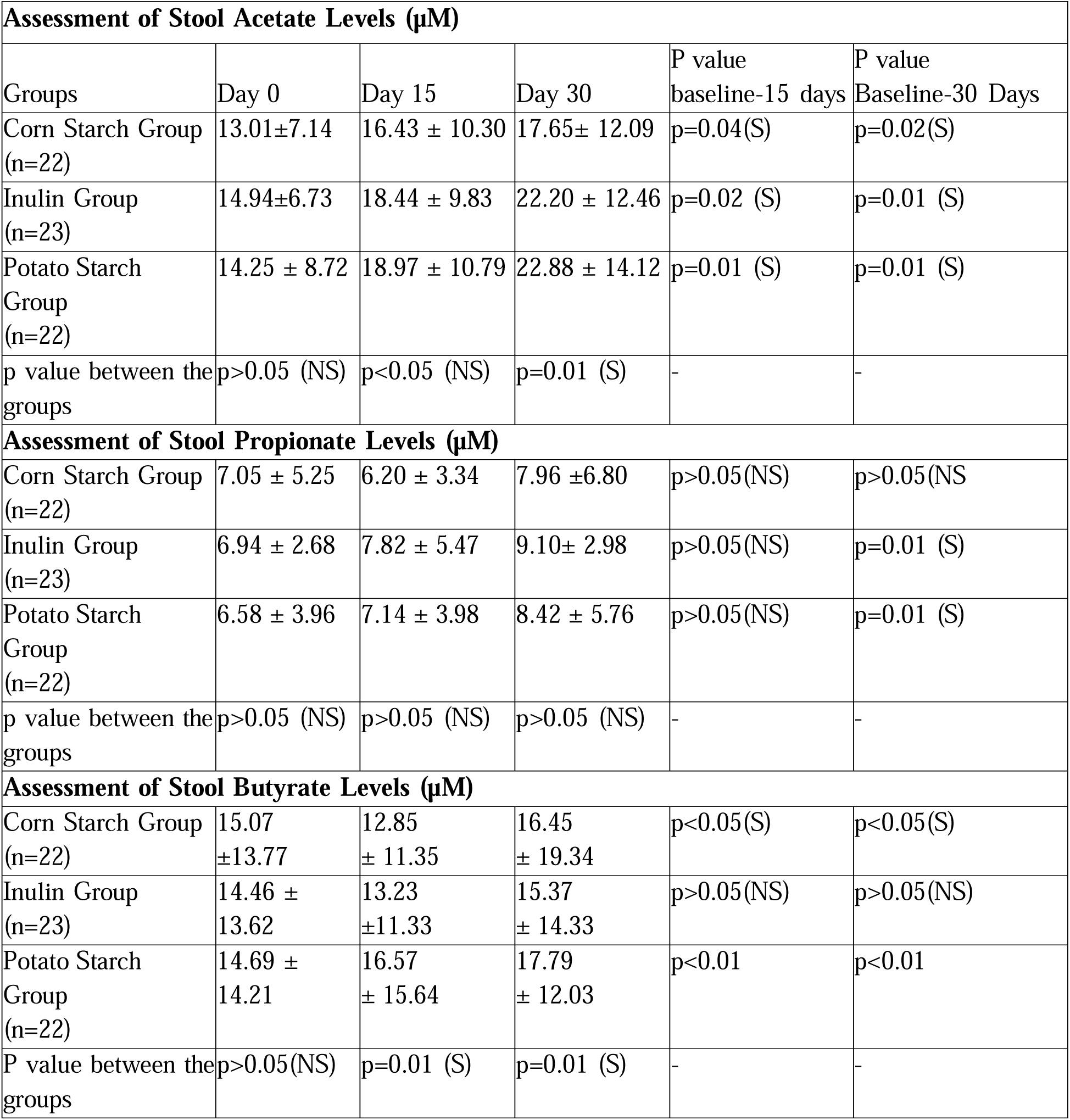
Assessment of Stool Acetate, Propionate and Butyrate Levels (µM)

| **Assessment of Stool Acetate Levels (μM)** |  |  |  |  |  |
| --- | --- | --- | --- | --- | --- |
| Groups | Day 0 | Day 15 | Day 30 | P value<br>baseline-15 days | P value<br>Baseline-30 Days |
| Corn Starch Group<br>(n=22) | 13.01±7.14 | 16.43 ± 10.30 | 17.65± 12.09 | p=0.04(S) | p=0.02(S) |
| Inulin Group<br>(n=23) | 14.94±6.73 | 18.44 ± 9.83 | 22.20 ± 12.46 | p=0.02 (S) | p=0.01 (S) |
| Potato Starch<br>Group<br>(n=22) | 14.25 ± 8.72 | 18.97 ± 10.79 | 22.88 ± 14.12 | p=0.01 (S) | p=0.01 (S) |
| p value between the<br>groups | p>0.05 (NS) | p<0.05 (NS) | p=0.01 (S) | - | - |
| **Assessment of Stool Propionate Levels (μM)** |  |  |  |  |  |
| Corn Starch Group<br>(n=22) | 7.05 ± 5.25 | 6.20 ± 3.34 | 7.96 ± 6.80 | p>0.05(NS) | p>0.05(NS) |
| Inulin Group<br>(n=23) | 6.94 ± 2.68 | 7.82 ± 5.47 | 9.10± 2.98 | p>0.05(NS) | p=0.01 (S) |
| Potato Starch<br>Group<br>(n=22) | 6.58 ± 3.96 | 7.14 ± 3.98 | 8.42 ± 5.76 | p>0.05(NS) | p=0.01 (S) |
| p value between the<br>groups | p>0.05 (NS) | p>0.05 (NS) | p>0.05 (NS) | - | - |
| **Assessment of Stool Butyrate Levels (μM)** |  |  |  |  |  |
| Corn Starch Group<br>(n=22) | 15.07<br>±13.77 | 12.85<br>± 11.35 | 16.45<br>± 19.34 | p<0.05(S) | p<0.05(S) |
| Inulin Group<br>(n=23) | 14.46 ±<br>13.62 | 13.23<br>±11.33 | 15.37<br>± 14.33 | p>0.05(NS) | p>0.05(NS) |
| Potato Starch<br>Group<br>(n=22) | 14.69 ±<br>14.21 | 16.57<br>± 15.64 | 17.79<br>± 12.03 | p<0.01 | p<0.01 |
| P value between the<br>groups | p>0.05(NS) | p=0.01 (S) | p=0.01 (S) | - | - |

#### Changes in stool propionate levels

It was observed that there was a significant reduction in the propionate levels from a baseline score of 7.05 ±5.25 to 6.20 ±3.34 in corn starch group at the end of 15 days. The levels however increased significantly to 7.96 ±6.80 at the end of 30 days. There was no significant change in stool propionate levels in the corn starch group by Day 30 (p>0.05). In the inulin group, the levels at baseline were 6.94 ±2.68 which increased significantly to 7.82 ±5.47 (p<0.05) and further to 9.10 ±2.98 (p=0.01) at the end of 15 days and 30 days respectively. The levels of stool propionate levels at baseline visit were 6.58 ±3.96 which showed a significant increase to 7.14 ±3.98 at the end of 15 days in the RPS group (p<0.05). The levels showed a further increase to 8.42 ±5.76 at the end of 30 days showing significant increase as compared to baseline (p=0.01). Analysis between the groups showed no significant difference between the groups. Percentage change in stool propionate levels from baseline to 15 days in Corn starch group were observed to be decreased by 12.05% while at 30 days it was observed to be increased by 12.90%. The change in stool propionate levels in the inulin group were observed to be 12.68% at 15 days and 31.12 % at the end of 30 days. The levels increased by 8.51 % at the end of 15 days and 27.96% at the end of 30 days in the RPS group. **Refer Table 3 & 4.**

**Table 4:** Percentage Change from Baseline in Stool Acetate, Propionate and Butyrate levels.

| **Percentage Change from Baseline in Stool Acetate levels** |  |  |
| --- | --- | --- |
| Study Groups | % Change Baseline – 15 Days | % Change Baseline – 30 Days |
| Corn Starch Group (n=22) | 26.28% | 35.66% |
| Inulin Group (n=23) | 23.42% | 48.59% |
| Potato Starch Group (n=22) | 33.12% | 60.56% |
| **Percentage Change from Baseline in Stool Propionate levels** |  |  |
| Corn Starch Group (n=22) | -12.05% | 12.90% |
| Inulin Group (n=23) | 12.68% | 31.12% |
| Potato Starch Group (n=22) | 8.51% | 27.96% |
| **Percentage Change from Baseline in Stool Butyrate levels** |  |  |
| Corn Starch Group (n=22) | -14.73% | 9.15% |
| Inulin Group (n=23) | -8.50% | 6.29% |
| Potato Starch Group (n=22) | 12.79% | 21.10% |

### Changes in stool butyrate levels

The baseline stool butyrate levels in corn starch group were observed to be 15.07 ±13.77 which showed a significant decrease to 12.85 ± 11.35 at the end of 15 days (p<0.05) and a significant increase to 16.45± 19.34 (p<0.05). The levels of stool butyrate levels in inulin group at baseline was observed to be 14.46 ± 13.62 which decreased non-significantly to 13.23 ±11.33 (p>0.05) at the end of 15 days and further increased non-significantly to 15.37 ± 14.33 (p>0.05) at the end of 30 days. In the RPS group, the stool butyrate levels at baseline visit was 14.69 ± 14.21 which showed a significant increase to a level to 16.57 ± 15.64 (p<0.01) at the end of 15 days and further to 17.79 ± 12.03 (p<0.01) at the end of 30 days. Analysis between the three groups showed that the increase in stool butyrate levels was significantly higher in RPS group as compared to inulin and corn starch groups. The concentration of butyrate in the RPS group was also significantly greater than that of the inulin and corn starch groups at both 15 days (p=0.01) and 30 days (p=0.01). The percentage change in stool butyrate levels from baseline to 15 days in Corn starch group were observed to have decreased by 14.73% while at 30 days, a net increase of 9.15% was observed. The change in stool butyrate levels in inulin group was observed to have decreased by 8.50% at 15 days and with a net increase of 6.29% at the end of 30 days. In the RPS group, butyrate concentration increased by 12.79% at the end of 15 days and 21.10% at the end of 30 days. **Refer Table 3 & 4.**

### Changes in stool SCFA levels (Total)

The baseline stool SCFA levels in the corn starch group was observed to be 35.12 ±20.29 which showed a slight non-significant increase to 35.48 ±14.35 at the end of 15 days and further significantly to 45.84 ±24.01 (p=0.02). The levels of stool SCFA levels in the inulin group at baseline was observed to be 36.33 ±14.34 which increased significantly to 39.49 ±13.45 (p=0.03) at the end of 15 days and further to 48.98 ±11.04 (p=0.01) at the end of 30 days. In the RPS group, the stool SCFA levels at baseline visit was 34.54 ±18.43 which showed a significant increase to 42.27 ±18.59 (p=0.01) at the end of 15 days and further to 49.18 ±15.46 at the end of 30 days (p=0.01). Analysis between the three groups showed that the increase in stool SCFA levels were significantly higher in the RPS group as compared to inulin and corn starch groups at 15 (p=0.01) and 30 days (p=0.04). Percentage changes in stool SCFA levels from baseline to 15 days in Corn starch group were observed to be 1.02% while at 30 days it was observed to be 30.52%. The change in stool SCFA levels in inulin group were observed to be 8.69% at 15 days and 34.81% at the end of 30 days. The levels increased by 14.64% at the end of 15 days and 42.38% at the end of 30 days in RPS group. **Refer Table 5 & 6.**

**Table 5:** Assessment of total SCFA levels in stool.

| **Assessment of total SCFA levels in stool** |  |  |  |  |  |
| --- | --- | --- | --- | --- | --- |
|  | **Day 0** | **Day 15** | **Day 30** | **P value**<br>**Baseline-15 days** | **P value –**<br>**Baseline-30 Days** |
| Corn Starch Group<br>(n=22) | 35.12<br>± 20.29 | 35.48<br>± 14.35 | 45.84<br>± 24.01 | p>0.05(NS) | p=0.02 (S) |
| Inulin Group<br>(n=23) | 36.33<br>±14.34 | 39.49<br>± 13.45 | 48.98<br>± 11.04 | p=0.03 (S) | p=0.01 (S) |
| Potato Starch Group<br>(n=22) | 34.54<br>±18.43 | 42.27<br>±18.59 | 49.18<br>±15.46 | p=0.01 (S) | p=0.01 (S) |
| P value between the<br>groups | p>0.05(NS) | p=0.01 (S) | p=0.04 (S) | - | - |

**Table 6:** Percentage change in SCFA levels in stool from baseline.

| **Study Groups** | **% Change Baseline – 15 Days** | **% Change Baseline – 30**<br>**Days** |
| --- | --- | --- |
| Corn Starch Group (n=22) | 1.02% | 30.52% |
| Inulin Group (n=23) | 8.69% | 34.81% |
| Potato Starch Group<br>(n=22) | 14.64% | 42.38% |

### Assessment of changes in Gut Microbiomes (diversity & abundance)

Gut microbiomes were analyzed specifically those producing butyrate. It was observed that there was a significant increase in the Bacteroidia and Actinobacteria levels. It was observed that there was a significant increase in the Bacteroidia, Firmicutes and Actinobacteria levels. **Refer Table 7 & 8. Figures 2,3,4 and 5.**

**Figure 2:**
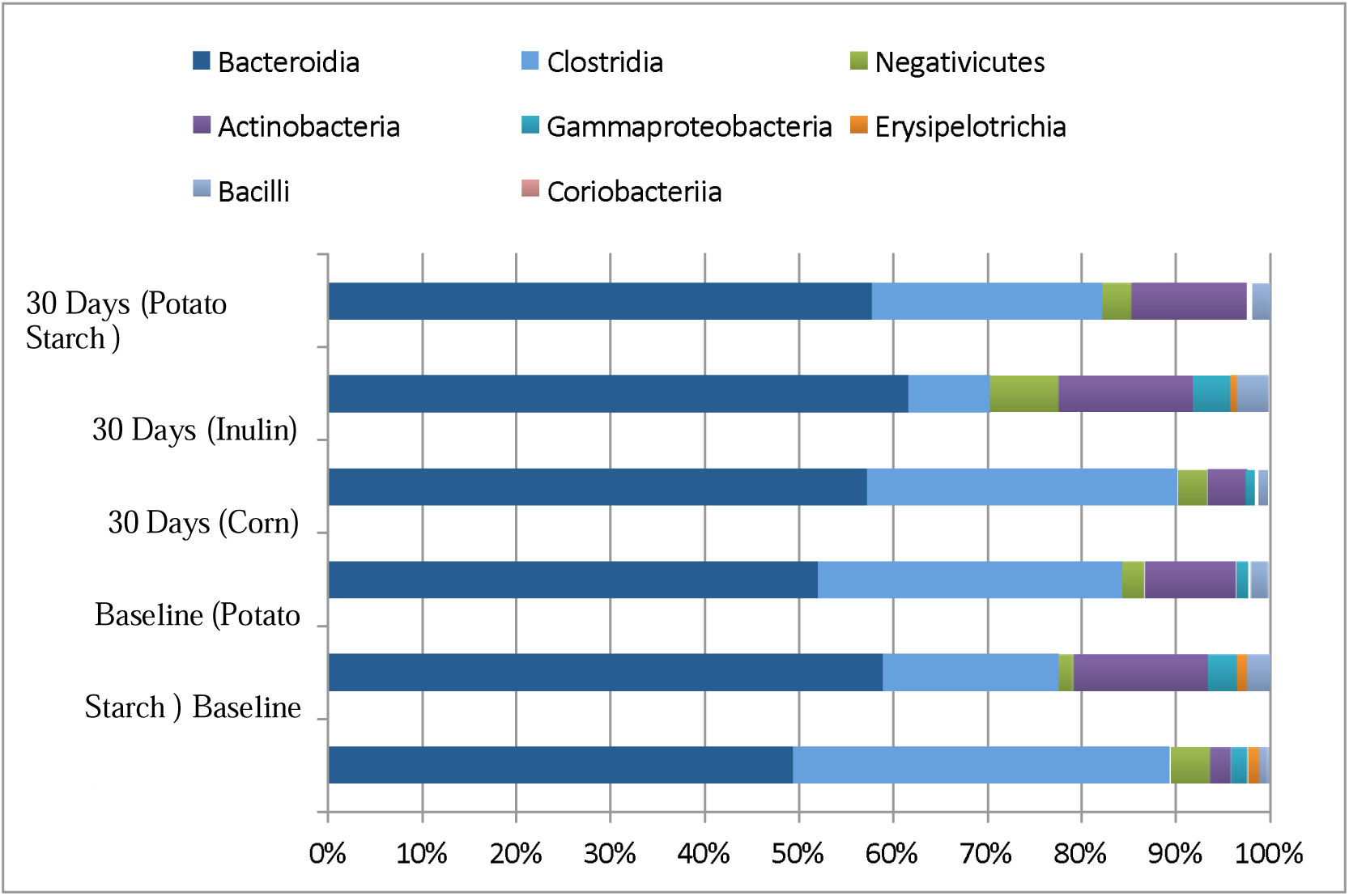
Abundance of SCFA (Specially butyrate) producing gut microbiome (Class) **Note**: Data represents Percentage on gut micro biomes (units 1=100%)

**FIG 3:**
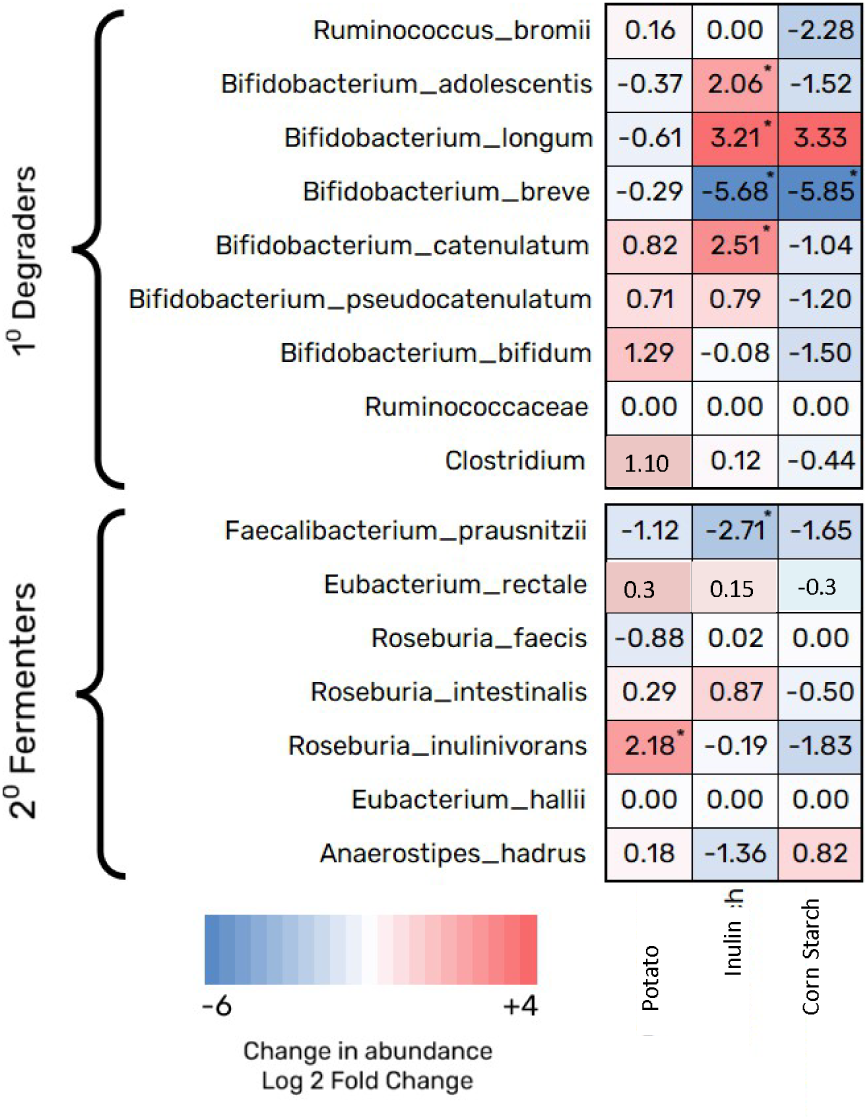
Average fold changes in the relative abundance of sequences representing selected primary (1°) degraders of resistant polysaccharides and secondary (2°) butyrate fermenters in response to dietary supplements

**FIG 4.**
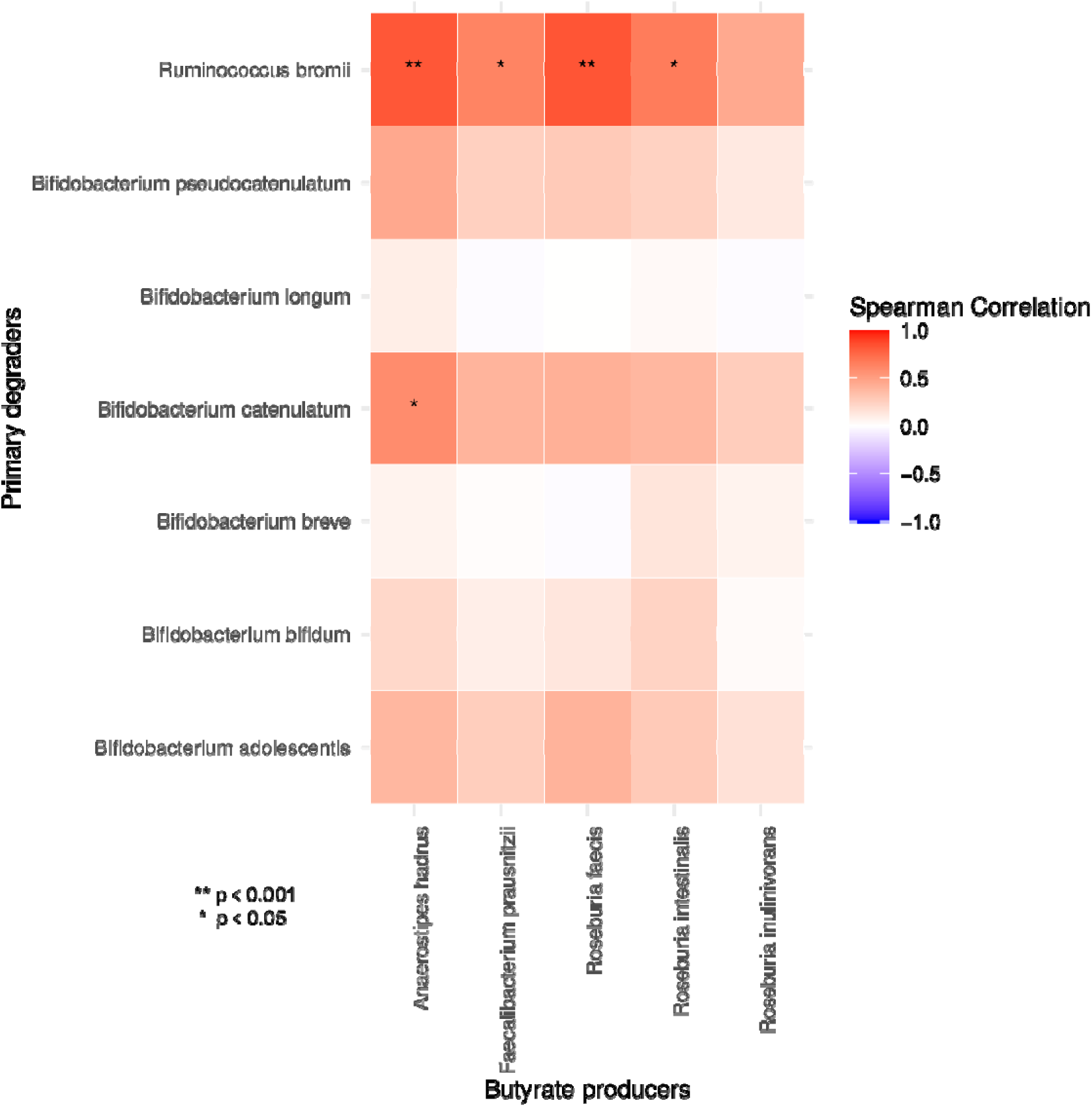
Pairs of microbes that consistently responded in concert either positively (red) or negatively (blue) to dietary supplementation with Resistant Potato Starch. Correlations between changes in the abundance of primary degraders and butyrate producers were calculated using the data set that includes responses with the use of RPS.

**Figure 5:**
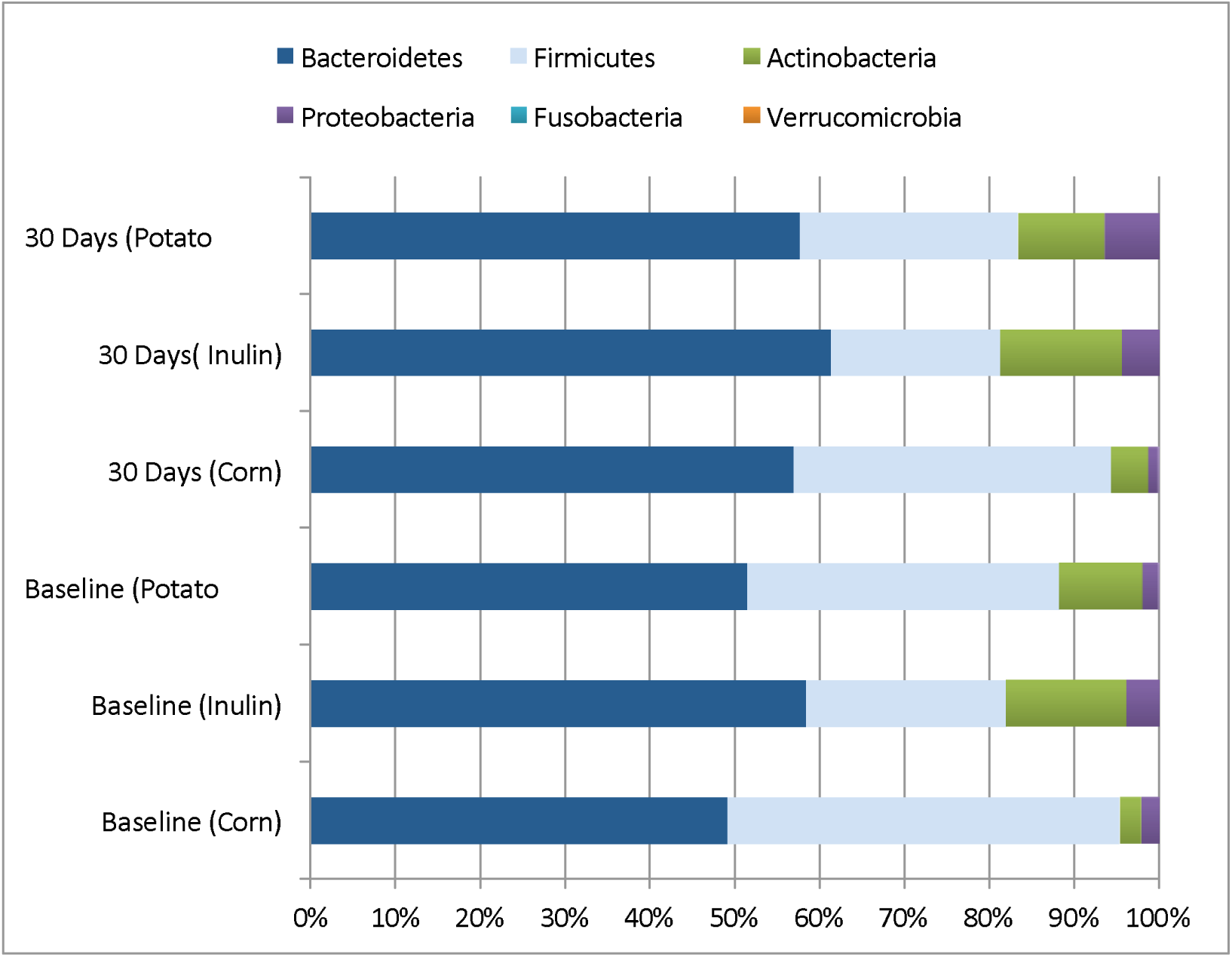
Abundance of SCFA (Specially butyrate) producing gut microbiome (Phylum) **Note**: Data represents Percentage on gut micro biomes (units 1=100%)

**Table 7:** Abundance of SCFA (Specially butyrate) producing gut microbiome (Class)

| **Gut Micro biome**<br>**(Class)** | **Baseline**<br>**(Corn)** | **Baseline**<br>**(Inulin)** | **Baseline**<br>**(Potato**<br>**Starch)** | **30 Days**<br>**(Corn)** | **30 Days**<br>**(Inulin)** | **30 Days**<br>**(Potatodaat)** |
| --- | --- | --- | --- | --- | --- | --- |
| Bacteroidia | 0.4912 | 0.5839 | 0.5146 | 0.5681 | 0.6132 | 0.5734 |
| Clostridia | 0.3984 | 0.1846 | 0.3194 | 0.3273 | 0.0862 | 0.2424 |
| Negativicutes | 0.0421 | 0.0159 | 0.0231 | 0.0318 | 0.0724 | 0.0305 |
| Actinobacteria | 0.0213 | 0.1411 | 0.0959 | 0.0401 | 0.1414 | 0.1212 |
| Gammaproteobacteria | 0.0184 | 0.0304 | 0.0102 | 0.0076 | 0.0401 | 0.0026 |
| Erysipelotrichia | 0.0110 | 0.0105 | 0.0052 | 0.0056 | 0.0069 | 0.0021 |
| Bacilli | 0.0094 | 0.0239 | 0.0187 | 0.0087 | 0.0334 | 0.0199 |
| Coriobacteriia | 0.0036 | 0.0007 | 0.0020 | 0.0041 | 0.0022 | 0.0003 |

**Note:** Data represents Percentage on gut microbiomes (units 1=100%)

**Table 8:**
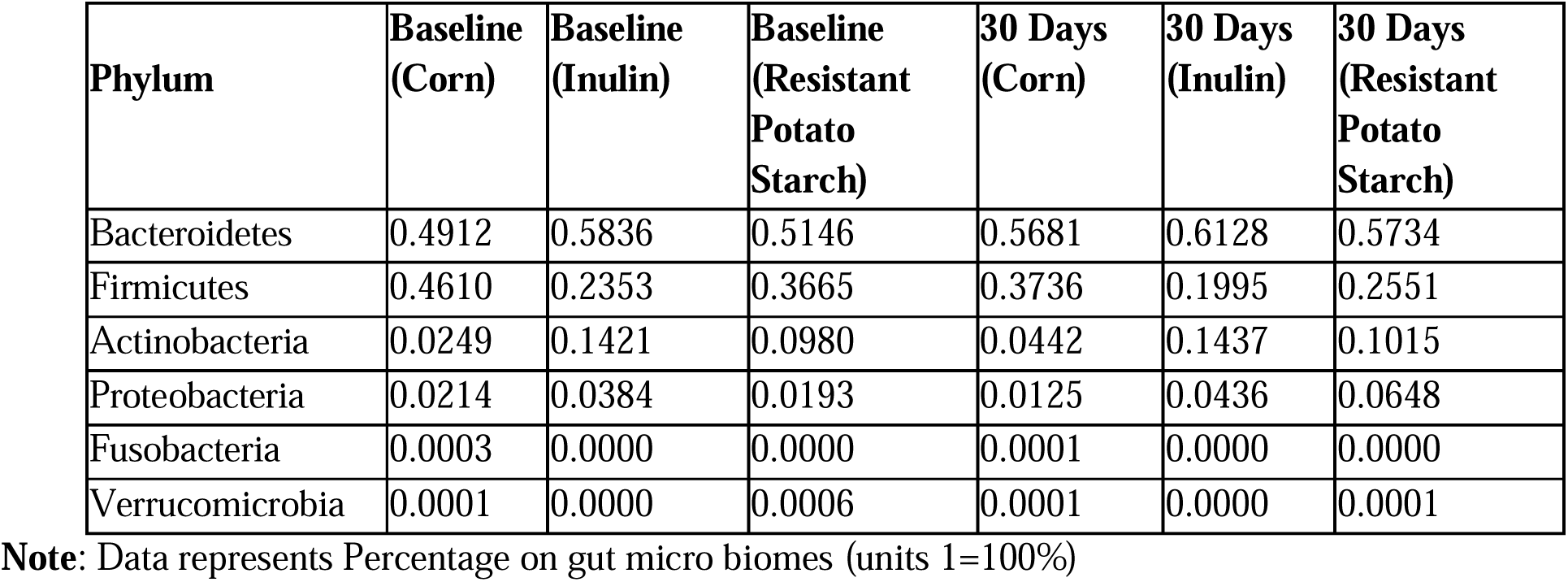
Abundance of SCFA (Specially butyrate) producing gut microbiome (Phylum)

| Phylum | Baseline (Corn) | Baseline (Inulin) | Baseline (Resistant Potato Starch) | 30 Days (Corn) | 30 Days (Inulin) | 30 Days (Resistant Potato Starch) |
| --- | --- | --- | --- | --- | --- | --- |
| Bacteroidetes | 0.4912 | 0.5836 | 0.5146 | 0.5681 | 0.6128 | 0.5734 |
| Firmicutes | 0.4610 | 0.2353 | 0.3665 | 0.3736 | 0.1995 | 0.2551 |
| Actinobacteria | 0.0249 | 0.1421 | 0.0980 | 0.0442 | 0.1437 | 0.1015 |
| Proteobacteria | 0.0214 | 0.0384 | 0.0193 | 0.0125 | 0.0436 | 0.0648 |
| Fusobacteria | 0.0003 | 0.0000 | 0.0000 | 0.0001 | 0.0000 | 0.0000 |
| Verrucomicrobia | 0.0001 | 0.0000 | 0.0006 | 0.0001 | 0.0000 | 0.0001 |

**Note:** Data represents Percentage on gut micro biomes (units 1=100%)

## Secondary outcomes

### Assessment of consistency of stool, digestion, appetite, bowel movements, abdominal fullness & flatulence

#### Change in stool consistency

Stool consistency was measured on Bristol scale. There was non-significant difference in the consistency of stool from baseline to 15 days and further to 30 days. The difference between the groups was also found to be non-significant. However, there was shift towards the ideal stool consistency in all three study groups. **Refer Table 9**.

**Table 9:** Assessment of stool on Bristol scale for consistency.

|  | **Day 0** | **Day 15** | **Day 30** | **P value**<br>**Baseline-15 days** | **P value –**<br>**Baseline-30 Days** |
| --- | --- | --- | --- | --- | --- |
| Corn Starch Group<br>(n=22) | 3.13 ±0.54 | 3.42 ±0.50 | 3.71 ±0.46 | p>0.05(NS) | p>0.05(NS) |
| Inulin Group<br>(n=23) | 3.39 ±0.58 | 3.61 ±0.50 | 3.61 ±0.51 | p>0.05(NS) | p>0.05(NS) |
| Resistant Potato<br>Starch Group<br>(n=22) | 3.43 ±0.59 | 3.61 ±0.50 | 3.77 ±0.43 | p>0.05(NS) | p>0.05(NS) |
| P value between the<br>groups | p>0.05(NS) | p>0.05 (NS) | p>0.05 (NS) | - | - |

#### Change in Digestion score

It was observed that there was a significant improvement in digestion from a baseline score of 70.21 ±14.63 to a final score of 83.54 ±9.26 showing an improvement of 18.98% in corn starch group. In the inulin group the score showed improvement from a baseline score of 67.39±16.23 to a score of 83.70±9.80 at the end of 30 days, showing an improvement of 24.20% over baseline score. In the RPS group, the score showed a significant improvement from a baseline score of 66.74±17.43 to a final score of 91.82±3.63 (p=0.04) at the end of the study, showing an improvement of 37.57% over baseline. On analysis between the groups, the change in digestion score was observed to be significantly better in the RPS group as compared to the other two study groups. **Refer Table 10**.

**Table 10:** Assessment of change in digestion.

|  | Corn starch | Inulin | Resistant<br>Potato Starch |
| --- | --- | --- | --- |
| Baseline | 70.21 ±14.63 | 67.39±16.23 | 66.74±17.43 |
| 15 Days | 77.71 ±10.00 | 79.13±10.19 | 79.13±9.00 |
| 30 Days | 83.54 ±9.26 | 83.70±9.80 | 91.82±3.63 |
| Change from baseline to 15 Days | 30.80% | 17.42% | 18.56% |
| Change from baseline to 30 Days | 18.98% | 24.20% | 37.57% |
| P value between the groups from<br>baseline to 30 days | p=0.04 (S) |  |  |

#### Change in Appetite Score

It was observed that there was a significant improvement in appetite from a baseline score of 69.38±12.96 to a final score of 83.54±8.40 showing a improvement of 23.29% in corn starch group. In the inulin group the score showed improvement from a baseline score of 69.78±14.50 to a score of 83.26±10.83 at the end of 30 days, showing an improvement of 19.31% over baseline score. In the RPS group the score showed a significant improvement from a baseline score of 70.43±10.65 to a final score of 91.36±3.63 at the end of the study, showing an improvement of 29.71% over baseline. On analysis between the groups the change in digestion score was observed to be significantly better in the RPS group as compared to the other two study groups. **Refer Table 11**.

**Table 11:** Assessment of change in appetite.

|  | Corn starch | Inulin | Resistant<br>Potato Starch |
| --- | --- | --- | --- |
| Baseline | 69.38±12.96 | 69.78±14.50 | 70.43±10.65 |
| 15 Days | 79.38±8.38 | 78.26±9.49 | 81.52±7.60 |
| 30 Days | 83.54±8.40 | 83.26±10.83 | 91.36±3.63 |
| Change from baseline to 15 Days | 14.41% | 12.15% | 15.74% |
| Change from baseline to 30 Days | 23.29% | 19.31% | 29.71% |
| P value between the groups from<br>baseline to 30 days | p=0.04 (S) |  |  |

#### Change in bowel habits Score

Bowel habits showed a significant improvement from base line score of 71.04±12.85 to a score of 82.71±10.32 at the end of 30 days with a 16.42% improvement. Inulin group, baseline score was 69.78±13.01 which improved significantly to 83.91±7.53 at the end of 30 days, showing an improvement of 20.24%. The RPS group showed an improvement in bowel habits score from a baseline of 69.57±11.74 to a score of 92.05±4.54 at the end of the study showing 32.31% improvement. Between group analysis showed statistically significant improvement in RPS group as compared to corn starch and inulin. **Refer Table 12**.

**Table 12:** Assessment of change in Bowel Habits.

|  | Corn starch | Inulin | Resistant<br>Potato Starch |
| --- | --- | --- | --- |
| Baseline | 71.04±12.85 | 69.78±13.01 | 69.57±11.74 |
| 15 Days | 79.79±11.28 | 81.52±7.30 | 80.00±8.66 |
| 30 Days | 82.71±10.32 | 83.91±7.53 | 92.05±4.54 |
| Change from baseline to 15 Days | 12.31% | 16.82% | 14.99% |
| Change from baseline to 30 Days | 16.42% | 20.24% | 32.31% |
| P value between the groups from baseline to 30 days | p=0.02 (S) |  |  |

#### Change in flatulence score

Flatulence reduced significantly with the use of RPS over a period of 30 days as compared to inulin and corn starch. At baseline visit the flatulence score was observed to be 18.44±14.57 which reduced by 24.45% to a score of 13.93±7.12 over 30 days in corn starch group. In the inulin group flatulence levels reduced from a score of 21.36±14.68 to a score of 10.91±7.01 at the end of 30 days showing a reduction of 48.92%. In the RPS group the scores showed a reduction of 63.72% from baseline score of 21.67±13.23 to final score of 7.86±5.67 at the end of 30 days. **Refer Table 13**.

**Table 13:**
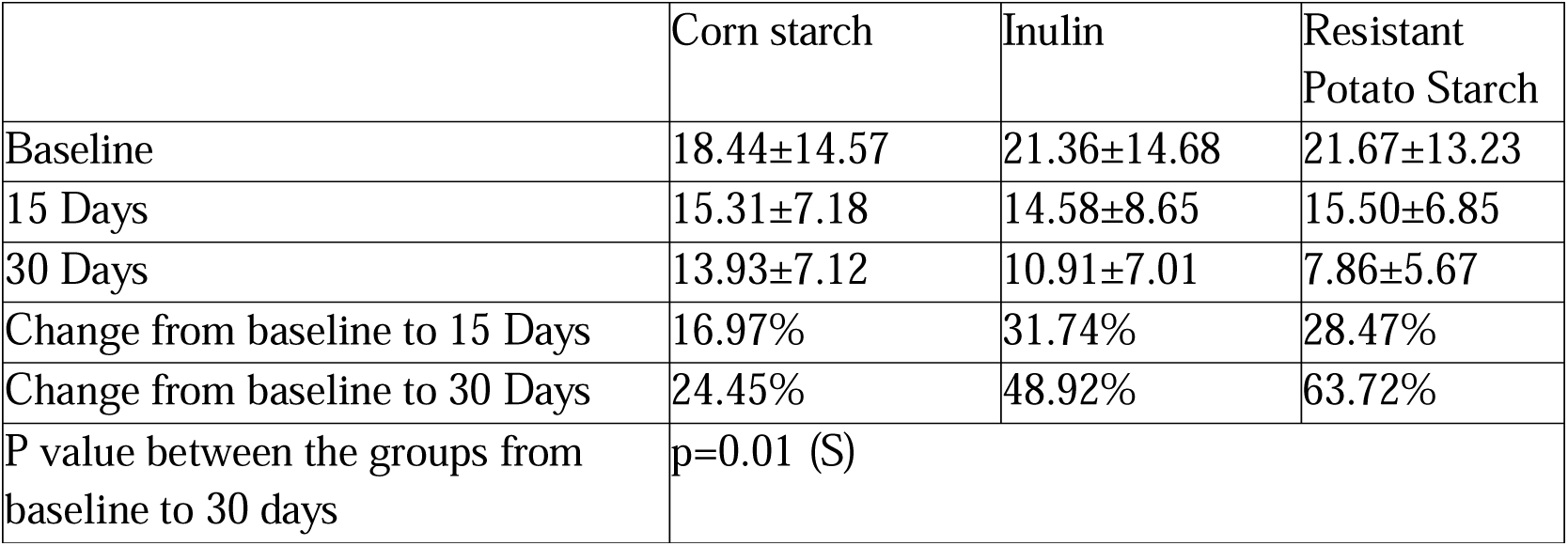
Assessment of change in Flatulence.

|  | Corn starch | Inulin | Resistant<br>Potato Starch |
| --- | --- | --- | --- |
| Baseline | 18.44±14.57 | 21.36±14.68 | 21.67±13.23 |
| 15 Days | 15.31±7.18 | 14.58±8.65 | 15.50±6.85 |
| 30 Days | 13.93±7.12 | 10.91±7.01 | 7.86±5.67 |
| Change from baseline to 15 Days | 16.97% | 31.74% | 28.47% |
| Change from baseline to 30 Days | 24.45% | 48.92% | 63.72% |
| P value between the groups from<br>baseline to 30 days | p=0.01 (S) |  |  |

#### Change in abdominal fullness score

Abdominal fullness reduced significantly with the use of RPS over a period of 30 days as compared to inulin and corn starch. At baseline visit the flatulence score was observed to be 14.00±7.37 which reduced by 30.78% to a score of 9.69±3.86 over 30 days in corn starch group. In the inulin group flatulence levels reduced from a score of 15.00±9.57 to a score of 9.55±4.16 at the end of 30 days showing a reduction of 36.33%. In the RPS group the scores showed a reduction of 68.85% from baseline score of 17.50±11.22 to final score of 5.45±1.51 at the end of 30 days. **Refer Table 14**.

**Table 14:** Assessment of change in Fullness of Abdomen.

|  | Corn starch | Inulin | Resistant<br>Potato Starch |
| --- | --- | --- | --- |
| Baseline | 14.00±7.37 | 15.00±9.57 | 17.50±11.22 |
| 15 Days | 11.25±3.87 | 11.54±5.16 | 11.07±5.25 |
| 30 Days | 9.69±3.86 | 9.55±4.16 | 5.45±1.51 |
| Change from baseline to 15 Days | 19.64% | 23.06% | 36.74% |
| Change from baseline to 30 Days | 30.78% | 36.33% | 68.85% |
| P value between the groups from<br>baseline to 30 days | p=0.01 (S) |  |  |

#### Global assessment of overall change, safety and adverse effects

It was observed that majority of participants were reported to having very much to much improvement. Also all the participants reported of excellent or good overall safety. There were 12 adverse events (3 in corn starch group, 4 in Inulin group and 4 in RPS group) recorded in the study which included fever, cough, cold, urine infection, small injuries. These AE were observed to be not related to any of the three study products.

## Discussion

This study investigated the efficacy of a new, proprietary resistant potato starch (Potatodaat^®^) as a prebiotic in comparison to inulin and an accessible corn starch control over a 30-day period. In this double-blinded, placebo-controlled study, it was found that the RPS group showed the most significant improvements in SCFA concentrations and clinical symptoms of IBS in comparison to inulin and accessible corn starch. The results of this study show the efficacy of this RPS as a dietary intervention and prebiotic.

This study built on previous studies that have examined the effect of resistant starches and prebiotics on the gut microbiome and short chain fatty acid concentrations at larger dosages. Dietary supplementation for 2 weeks with 28-34 g/day of RPS has been shown to have a greater butyrogenic effect than resistant maize starch, inulin, or accessible corn starch. This study shows this effect is replicable at lower dosages and with longer supplementation time.

Prebiotics significantly influence the composition and activity of the gut microbiome 54.Naseer M., Poola S., Uraz S., Tahan V. Therapeutic Effects of Prebiotics on Constipation: A Schematic Review. Curr. Clin. Pharmacol. 2020;15:207–215. doi: 10.2174/1574884715666200212125035. Prebiotics promote the proliferation of beneficial bacteria such as *Bifidobacterium* and *Lactobacillus*, which are known for their positive effects on gut health Kleerebezem M., Vaughan E.E. Probiotic and gut lactobacilli and bifidobacteria: Molecular approaches to study diversity and activity. Annu. Rev. Microbiol. 2009;63:269–290. doi: 10.1146/annurev.micro.091208.073341. This shift in microbial composition can suppress the growth of harmful bacteria like *Clostridium perfringens* and *Escherichia coli*, thereby reducing the risk of gastrointestinal infections and inflammation. Smolinska S, Popescu FD, Zemelka-Wiacek M. A Review of the Influence of Prebiotics, Probiotics, Synbiotics, and Postbiotics on the Human Gut Microbiome and Intestinal Integrity. J Clin Med. 2025 May 23;14(11):3673. doi: 10.3390/jcm14113673. PMID: 40507435; PMCID: PMC12156228.

The greater improvement in digestive health that was seen in the RPS group relative to the inulin and placebo groups is supported by the analysis of the gut microbiome. The resistant starch acts as food for fermenting bacteria which produce intermediates that feed butyrate-producing bacteria. Changes in the gut microbiome of study participants supplemented with RPS was marked by increases in key bacteria shown to support butyrogenic activity through cross-feeding or direct production of butyrate. Overall gut microbiome analysis by phyla showed an increase in 3 phyla involved in starch fermentation: Firmicutes, Bacteroidetes, and Actinobacterium. Within the Firmicutes group, several species of R. bromii and Clostridium were specifically identified as resistant polysaccharides primary degraders that increased in abundance due to RPS supplementation. Similarly, several resistant polysaccharide primary degrading species of Bifidobacterium from the Actinobacterium phyla, including B. catenulatum, B. pseudocatenulatum, and B. bifidum, were also increased.

The butyrate producers that increased due to RPS supplementation included E. rectale, Roseburia intestinalis, Roseburia inulinivorans, and A. hadrus. Several bacteria from this group are from the clostridial cluster XIVa, known for butyrate-production. Additionally, the RPS group showed significant response in pairs of bacteria that cross-feed to enhance butyrate production, indicating that abundance was correlated between the pairs. These pairs include the degrader, R. bromii with the butyrate-producing bacterial species, A. hadrus, F. prausnitzii, Roseburia faecus, and Roseburia intestinalis.

The changes seen in the gut microbiome analysis validate the increased butyrate and SCFA concentration from the stool analysis. The significant increase in the acetate intermediates seen in the SCFA panel of the RPS group indicates it produced the most change in the concentration for cross-feeding of the butyrogenic bacteria. Consequently, in comparison to inulin and placebo, the RPS group also showed the greatest butyrogenic activity through the largest increase in butyrate concentration.

While the direct measure of butyrate showed that RPS had the greatest impact on the gut microbiome, this activity also translated to clinical, digestive benefits. The secondary clinical assessments showed the efficacy of dietary intervention with RPS in comparison to the inulin and accessible corn starch. The RPS group showed the greatest improvement in digestion, appetite, bowel habits, flatulence, and abdominal fullness.

The modern Western diet lacks in dietary fiber which contributes to the use prebiotic fibers as dietary supplements to improve digestive health, including inulin, fructo-oligosaccharides, and galacto-oligosaccharides. However, excessive use of these products can lead to negative side effects such as bloating, gas, and diarrhea. Inulin, a commonly used prebiotic though have been extensively used but is known to cause Nausea, bloating, and flatulence specially in people suffering from inflammatory bowel disease (IBD) or allergies. Sheng W, Ji G, Zhang L. Immunomodulatory effects of inulin and its intestinal metabolites. Front Immunol. 2023 Aug 10;14:1224092. doi: 10.3389/fimmu.2023.1224092. PMID: 37638034; PMCID: PMC10449545. Resistant starch, specifically resistant potato starch, has the potential to be a clean label alternative to these products without the negative side effects found in these alternative sources of fiber.

The three-tiered analysis in this study comparing benefits of RPS on a microbiological, metabolic, and clinical level supports the claim that this resistant potato starch functions as a more efficacious alternative to inulin as a prebiotic to support digestive health.

Limitations of this study include a limited sample size in each study arm and participants being focused on one geographic region. While assessments were completed to ensure subjects had a varied diet, alternative sources of fiber in the diet were not standardized. Additional studies that look at long-term benefits of this intervention as well as benefits from smaller doses of the intervention are warranted.

## Conclusion

The present study concludes that resistant potato starch was effective in significantly increasing stool short chain fatty acids including stool butyrate and stool acetate levels as compared to corn starch and inulin over a period of 30 days of use. There was also a significant improvement in digestive functions including digestion, appetite, bowel habits and decrease in flatulence and abdominal fullness. Effect on gut microbiota also showed significant increase in abundance and diversity of protective gut microbiomes. There was also a significant increase in the levels of total SCFA levels. These activities signify higher prebiotic activity of resistant potato starch as compared to corn starch and inulin. There were no adverse effects reported with the consumption of resistant potato starch.

## Data Availability

All data produced in the present work are contained in the manuscript

## Notes

### Competing Interest Statement

The authors have declared no competing interest.

### Clinical Trial

CTRI/2022/12/048064
[Registered on: 13/12/2022]

### Author Declarations

Independent Ethics Committee, Mhaske Hospital and Research Centre, Hadapsar, Pune. Independent Ethics Committee, Mhaske Hospital andResearch Centre, Hadapsar, Pune. DR DYPCA & RC INSITUTIONAL ETHICS COMMITTEE PIMPRI PUNE -18

